# Contribution of an under-recognized adversity to child health risk: large-scale, population-based ACEs screening

**DOI:** 10.1101/2025.02.04.25321682

**Authors:** Laura M. Glynn, Sabrina R. Liu, Charles Golden, Michael Weiss, Candice Taylor Lucas, Dan M. Cooper, Louis Ehwerhemuepha, Hal S. Stern, Tallie Z. Baram

## Abstract

**Background and Objectives:** Whereas adverse early life experiences (ACEs) correlate with cognitive, emotional and physical health at the population level, existing ACEs screens are only weakly predictive of outcomes for an individual child. This raises the possibility that important elements of the early-life experiences that drive vulnerability and resilience are not being captured. We previously demonstrated that unpredictable parental and household signals constitute an ACE with cross-cultural relevance. We created the 5-item Questionnaire of Unpredictability in Childhood (QUIC-5) that can be readily administered in pediatric clinics. Here, we tested if combined screening with the QUIC-5 and an ACEs measure in this real-world setting significantly improved prediction of child health outcomes.

**Methods:** Leveraging existing screening with the Pediatric ACEs and Related Life Events Screener (PEARLS) at annual well-child visits, we implemented QUIC-5 screening in 19 pediatric clinics spanning the diverse sociodemographic constituency of Orange County, CA. Children (12yr+) and caregivers (for children 0-17years) completed both screens. Health diagnoses were abstracted from electronic health records (N=29,305 children).

**Results:** For both screeners, increasing exposures were associated with a higher probability of a mental (ADHD, anxiety, depression, externalizing problems, sleep disorder) or physical (obesity abdominal pain, asthma, headache) health diagnosis. Across most diagnoses, PEARLS and QUIC provided unique predictive contributions. Importantly, for three outcomes (depression, obesity, sleep disorders) QUIC-5 identified vulnerable individuals that were missed by PEARLS alone.

**Conclusions:** Screening for unpredictability as an additional ACE in primary care is feasible, acceptable and provides unique, actionable information about child psychopathology and physical health.

**What’s Known on This Subject:** Whereas ACEs correlate with neurodevelopmental and physical health of children at the population level, ACEs scales (e.g., PEARLS) are only weakly predictive at the level of the individual child. Are important elements of early-life adversity missed by these scales?

**What This Study Adds:** Because unpredictable signals constitute a unique ACE, we developed the Questionnaire of Unpredictability in Childhood (QUIC-5). Administering QUIC-5 and PEARLS to 30,000 families identified youth at risk for depression, obesity and other health problems, who would be missed by PEARLS alone.

## Introduction

The Centers for Disease Control and Prevention-Kaiser Permanente study^1^ focusing on Adverse Childhood Experiences (ACEs) documented cumulative effects of exposure to potentially traumatic experiences (e.g. abuse, neglect, violence exposure) on a wide range of physical and mental health conditions. This seminal work prompted studies examining the enduring role of ACEs in health and disease^2^ and it is now estimated the economic burden of exposure to ACEs in the US adult population is $14.1 trillion annually^3^. Given the accumulating evidence regarding the human and fiscal toll of ACEs, calls to address the prevalence and consequences of ACEs are rapidly increasing^4,5^ and the vast majority of states have enacted some form of legislation to address the burden of ACEs^6^. At the forefront of these efforts, in 2020, California became the first state to implement a publicly supported screening program comprising guidelines for trauma-informed care coupled with reimbursement for ACEs screening for the State’s 15 million individuals supported by Medicaid. While California intends to dramatically reduce the burden of ACEs on its citizens through this program^7,8^, the value of this public health initiative has been questioned on several grounds^9^. First, whereas ACEs screens identify risk at the population level, they are limited in the ability to do so at the level of the individual^10^. In addition, in the absence of established means for the prevention or mitigation of the effects of ACEs, the value of screening is unclear^11^.

There are several potential explanations for why ACEs scores are limited in their ability to detect health risk for an individual child. One possibility is that significant sources of stress and trauma occurring in childhood are missed with existing screeners^12^. One such early life exposure that is not currently included in standard screening instruments is unpredictability of the family and environmental signals received by the child, which activate the brain’s stress responses. It has now been demonstrated in prospective longitudinal studies across diverse cultures and sociodemographic groups that unpredictable parental care (independent from parental support and sensitivity) and lack of structure in the family and home environment strongly predict cognitive and emotional development^13^. Notably, the concept that unpredictable signals to the developing brain disrupt brain maturation is well supported by experimental animal studies^14–17^. Specifically, unpredictability in childhood has been linked to decreased self-regulation, a slower trajectory of cognitive development and poorer memory, as well as increased risk for anxiety, depression, anhedonia, PTSD, and poorer self-reported physical health in children and adults^18–22^. These associations persist after consideration of other well-established ACEs (e.g. poverty, abuse, neglect), suggesting that unpredictable experience is a robust risk factor for adverse developmental and health outcomes, and its absence from existing assessments of early-life adversity may account for some of their shortcomings in predicting an individual child’s risk profile.

Here we test the relative and cumulative contributions of both ACES and unpredictability as risk factors for child mental and physical health in a large, diverse pediatric population. We leverage the existing ACEs screening implemented in the Children’s Hospital of Orange County (CHOC) primary care network^23^ together with a well-validated 5-item screening instrument for unpredictability to address the following critical questions: 1. When employed in routine pediatric primary care, does ACEs screening identify children at increased risk of mental and physical health problems? 2. Does screening for unpredictability in the home environment provide additional predictive power to current ACEs screening recommendations?

## Methods

### Study Setting and Participants

This study took place in 19 pediatric primary care clinics affiliated with Children’s Hospital of Orange County (CHOC), which serve a diverse community of children in Orange County, CA (see Table 1 for an overview of demographics). In 2020, the primary care clinics implemented routine ACEs screening for all children at their annual well-child visits. As part of the California Initiative to Advance Precision Medicine^24^, in 2021, optional screening for unpredictability was initiated. For both screens all caregivers provided information and the screens were also administered to children aged 12 and over. Inclusion criteria for the current study included: 1. Completion of the QUIC-5 screener. 2. Child ages 0 to 17 years. 3. English or Spanish language preference. Here we present data for the first 29,305 children screened for ACEs and unpredictability. All study procedures were approved by the CHOC institutional review board.

**Table 1.** Participant Characteristics (N = 29,305).

|  | **n** | **%** |
| --- | --- | --- |
| **Child Age** |  |  |
| *0-4* | 5,833 | 19.9 |
| *5-9* | 7,909 | 30.0 |
| *10-14* | 9,278 | 31.7 |
| *15-17* | 6,285 | 21.4 |
| **Child Race** |  |  |
| *Asian* | 2,892 | 9.9 |
| *Black* | 506 | 1.7 |
| *Native American or Alaskan* | 114 | 0.4 |
| *Native Hawaiian* | 22 | 0.1 |
| *White* | 7,400 | 25.3 |
| *More than one race or other* | 9,219 | 30.4 |
| *Unknown* | 9,152 | 31.2 |
| **Child Ethnicity** |  |  |
| *Hispanic/Latino/a* | 17,472 | 58.5 |
| *Not Hispanic/Latino/a* | 8,903 | 29.8 |
| *Unknown* | 3,486 | 11.7 |
| **Preferred Language** |  |  |
| *English* | 24,263 | 82.8 |
| *Spanish* | 5,042 | 17.2 |
| **Child Gender** |  |  |
| *Female* | 14,169 | 48.4 |
| *Male* | 15,136 | 51.6 |
| **Insurance** |  |  |
| *Public* | 20,112 | 68.7 |
| *Private* | 9,041 | 30.9 |
| *Self or Other* | 152 | 0.5 |

Note: Demographic data were retrieved from electronic health records. Race, ethnicity, and gender were collected from the caregiver at initial registration. Preferred language indicates the language in which the screens were administered. Insurance status was determined from the visit concurrent with screening.

### Assessment of ACEs

ACEs were assessed with the face-valid Pediatric and Related Life Events Scale (PEARLS^25^). Following current state screening recommendations, we examined the score for Part 1, which focuses on 10 ACEs yielding a potential score of 0 to 10. Current protocols at the CHOC Pediatric Primary care clinics utilize the aggregated or deidentified version of the PEARLS in which the respondent provides a count of the number of items positively endorsed without specification of the individual items contributing to these scores. PEARLS scores are available in electronic health records (EHR).

### Assessment of Unpredictability

Unpredictability was assessed with the 5-item version of the Questionnaire of Unpredictability in Childhood (QUIC-5^20,26^). At the well-child visit, caregiver and child (as appropriate based on age) were given the opportunity to complete the unpredictability screen. The QUIC, which broadly assesses unpredictability in the social, emotional and physical environments demonstrates robust psychometric properties. The scale was validated against prospective longitudinal assessments of early life unpredictability and exhibits strong content and discriminant validity as well as excellent test-retest reliability^20^. The QUIC-5, on which scores range from 0 to 5, is correlated on average .84 with the full-length version and predicts mental health outcomes effects sizes comparable to the original scale^26^.

### Child Health

Mental and Physical health conditions were selected based on those that have strong stress-related and behavioral components and have been previously identified as common outcomes of exposure to early life adversity in pediatric populations^27^. Presence or absence of the following conditions in each child’s EHR prior to or concurrent with the well-child visit at which the screen was conducted were obtained: abdominal pain, ADHD, anxiety, asthma, depression, externalizing problems, headache, obesity and sleep disorders. Specific ICD-10 codes for each diagnosis can be seen in Table S1 and the incidence of each diagnosis in Table S2.

### Analysis Plan

First, distributions of caregiver and youth endorsements on the PEARLS and QUIC were examined. Bivariate correlations were then used to determine the degree of association between youth and caregiver reports for each screener, as well as the correlation between the PEARLS and QUIC-5. For both QUIC-5 and PEARLS separate binary logistic regressions were conducted to examine the associations between caregiver and youth reports with mental and physical health outcomes. In these regressions, scores on both screeners were categorized as 0, 1, 2, 3 and 4+. To determine whether the QUIC adds predictive power to the PEARLS, we employed two approaches: 1. QUIC and PEARLS scores (continuous) were entered simultaneously into binary logistic regressions predicting mental and physical health outcomes.

1. 2. In a second set of binary logistic regressions, we examined the independent contributions of all possible combinations of QUIC (0, 1, 2, 3 and 4+) and PEARLS scores (0, 1, 2, 3 and 4+) with 0-0 as the reference for mental and physical health outcomes. All regression models adjusted for child gender and age (with both linear and quadratic age terms considered as appropriate).

## Results

### Exposures to ACEs and Unpredictability

Figure 1 shows the distribution of caregiver endorsement for exposures on PEARLS and QUIC among children ages 0 to 17 years. On average older children experienced more exposures to both ACEs and unpredictability, as expected. However, exposures to at least one ACE (13%) or at least one form of unpredictability (23%) were observed even among the youngest children ages 0-4 (means and standard deviations by age group are provided in Table S3).

**Figure 1.**
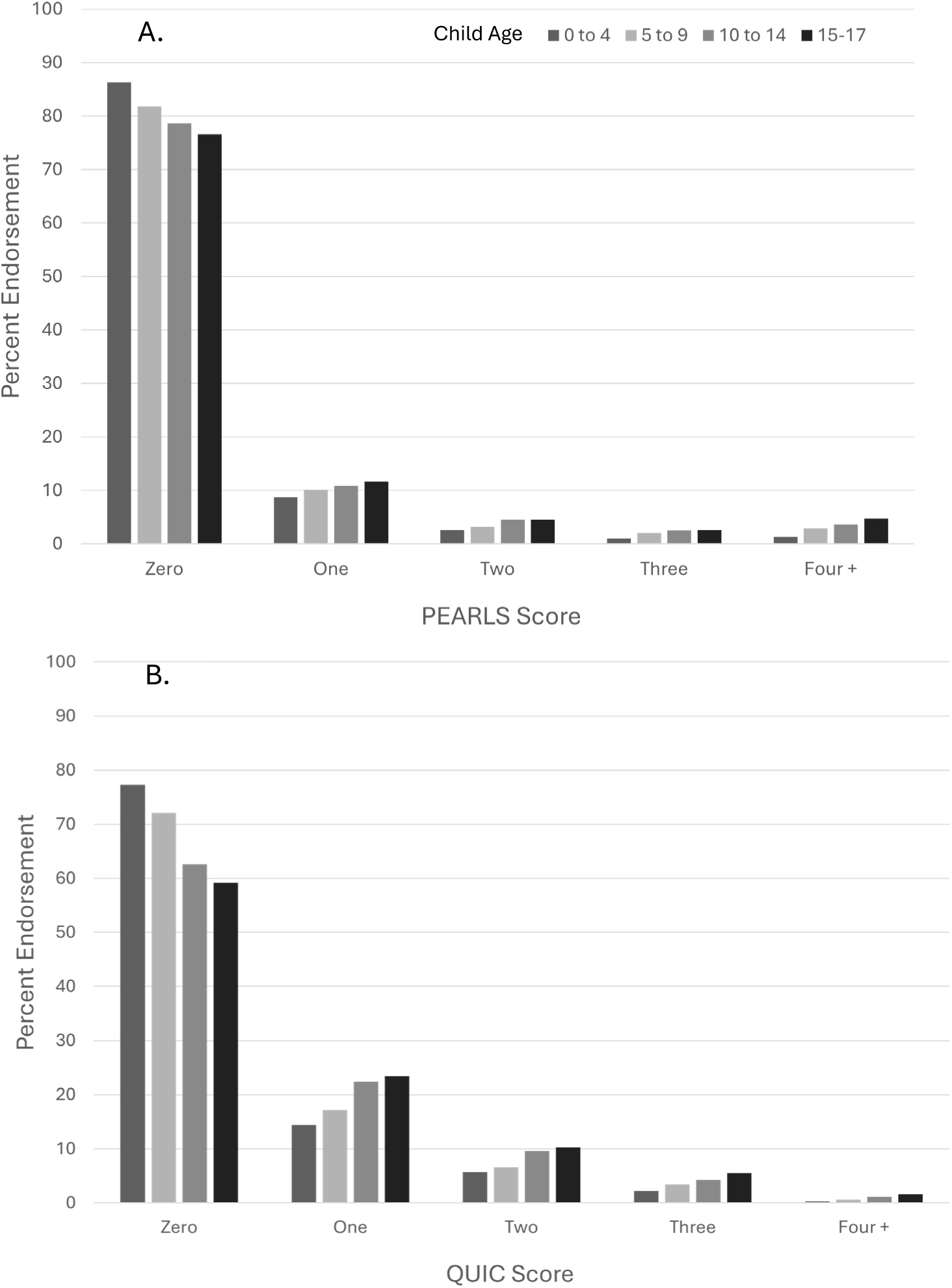
Distribution of ACEs (PEARLS) and unpredictability (QUIC-5) by child age.

Comparing self-report by the children to caregiver reports within the same dyads, youths reported more exposures to both ACEs and unpredictability than did caregivers (Figure 2; means and standard deviations provided in Table S4). The correlation between youth and caregiver reports on the PEARLS was .64 and for the QUIC the association was .54 (both p’s = .00). The two scales were also associated: among youth, the correlation between the QUIC-5 and PEARLS was .51, and the correlation of caregiver reports on the two screeners was of a similar magnitude (r = .43; both p’s = .00).

**Figure 2.**
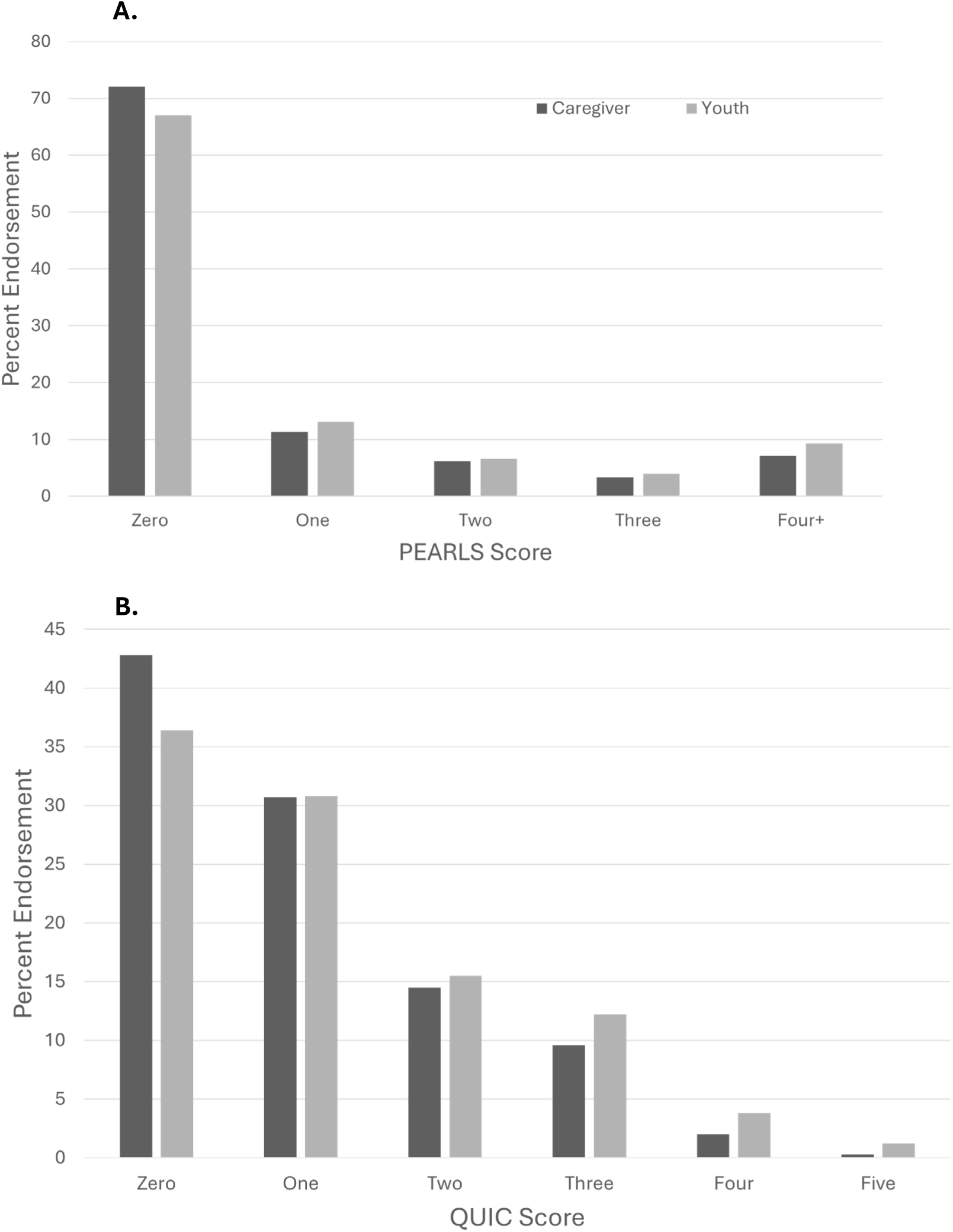
Distribution of ACEs (PEARLS; Panel A) and unpredictability (QUIC-5; Panel B) by caregiver report and youth self-report.

### Exposures To ACEs and Unpredictability Predict Child Mental and Physical Health

For both ACEs assessed with PEARLS and unpredictability measured with QUIC-5, increasing exposures were associated with a higher probability of a mental (ADHD, anxiety, depression, externalizing problems, sleep disorder) or physical (abdominal pain, asthma, headache, obesity) health diagnosis. This was true for reports by both caregiver and youth (Figures 3 and 4). Across diagnoses, the associations were generally dose-dependent, with odds ratios increasing with each additional exposure (ORs and CIs are provided in Tables S5-8). For caregiver report of mental health outcomes, the odds ratios for those with 4 or more exposures compared to those with zero ranged from 2.4 to 5.9 for the PEARLS and 1.9 to 3.4 for the QUIC. Similarly, the odds ratios for the physical health outcomes ranged from 1.4 to 1.9 and 1.8 to 2.0 for PEARLS and QUIC, respectively. The range of odds ratios for the youth self-reports were similar in magnitude (see Table S8).

**Figure 3.**
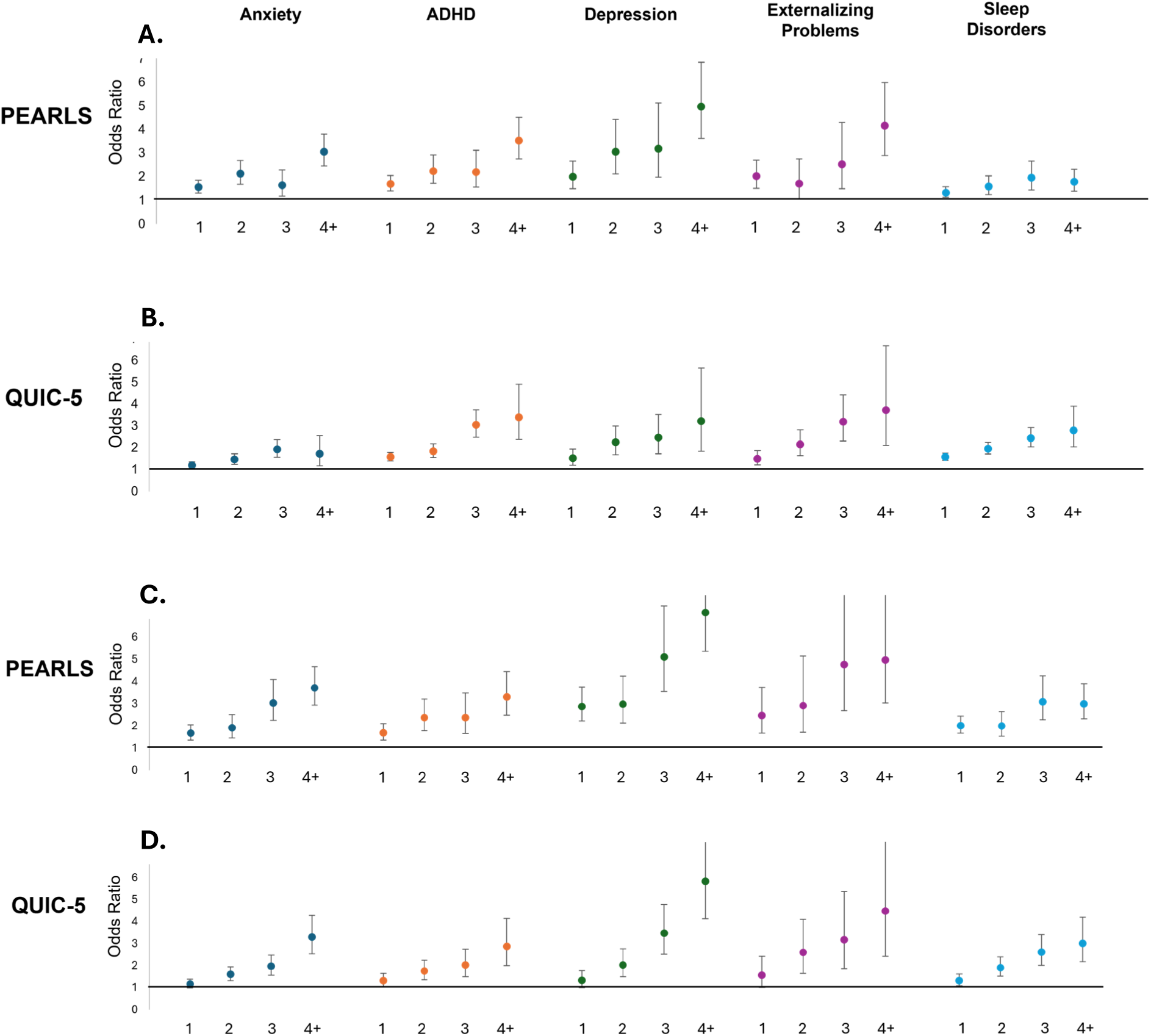
ACEs (PEARLS) and unpredictability (QUIC-5) screens predict child mental health outcomes. Odds ratios and 95% confidence intervals. Panels A-B show associations for caregiver reports. Youth self-report in panels C-D.

**Figure 4.**
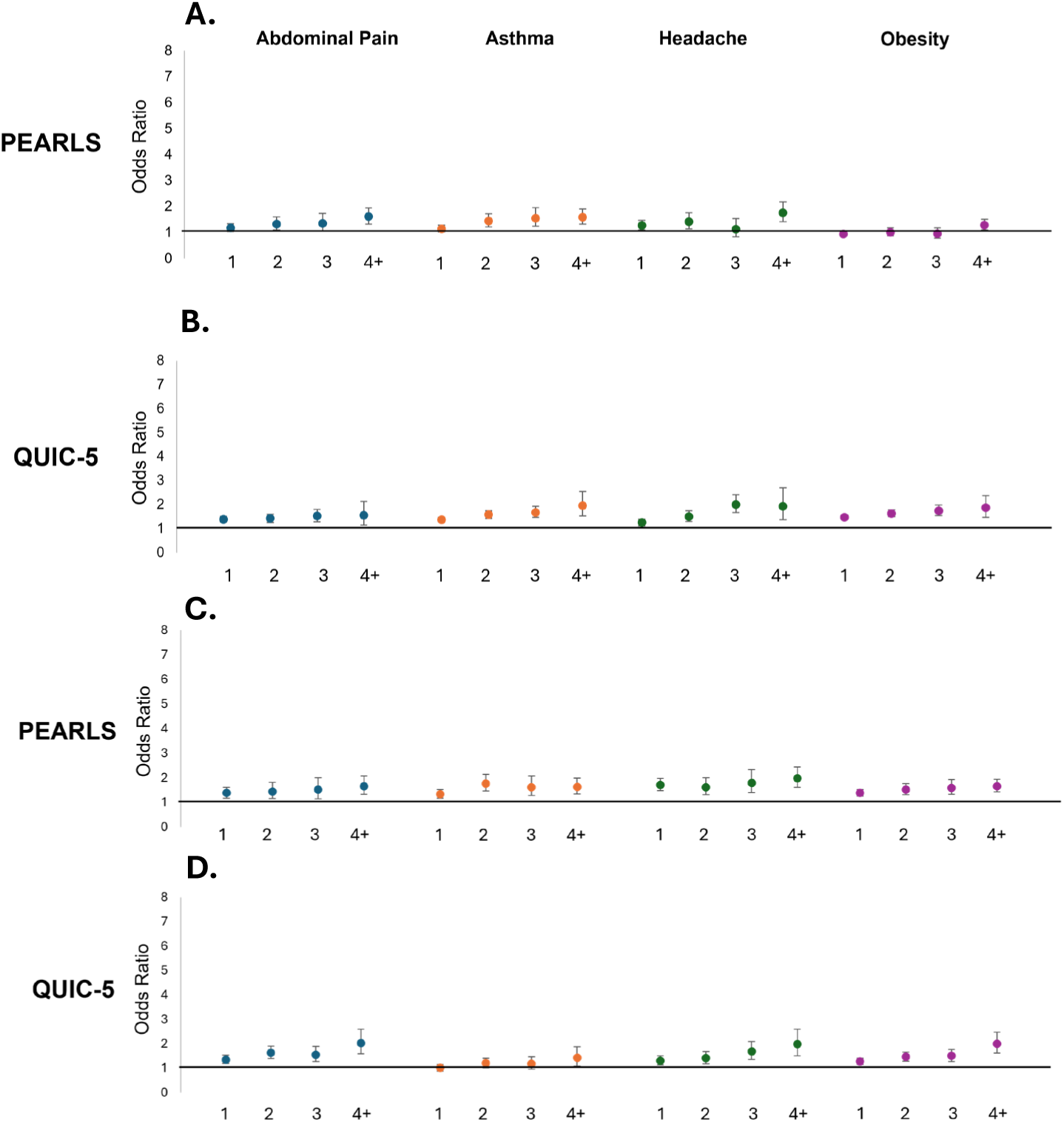
ACEs (PEARLS) and unpredictability (QUIC-5) screens predict child physical health outcomes. Odds ratios and 95% confidence intervals. Panels A-B show associations for caregiver reports. Youth self-report in panels C-D.

### Testing the Added Value of Unpredictability

The correlation between PEARLS and QUIC-5 scores raised the question of whether including the assessment of unpredictability adds value for predicting child health outcomes beyond that obtained for PEARLS alone. Therefore, we determined the adjusted ORs for both the QUIC and PEARLS in logistic regressions in which continuous scores for both were concurrently included as predictors of youth mental and physical health (Figure 5; Tables S9 and S10). For most mental and physical health diagnoses examined, both the PEARLS and QUIC provided unique predictive contributions, and the predictive power (odds ratios) were similar, whether reported by caregiver or youth. For sleep disorders and obesity the QUIC was a stronger predictor of the increased probability of a diagnosis than was the PEARLS (Figure 5).

**Figure 5.**
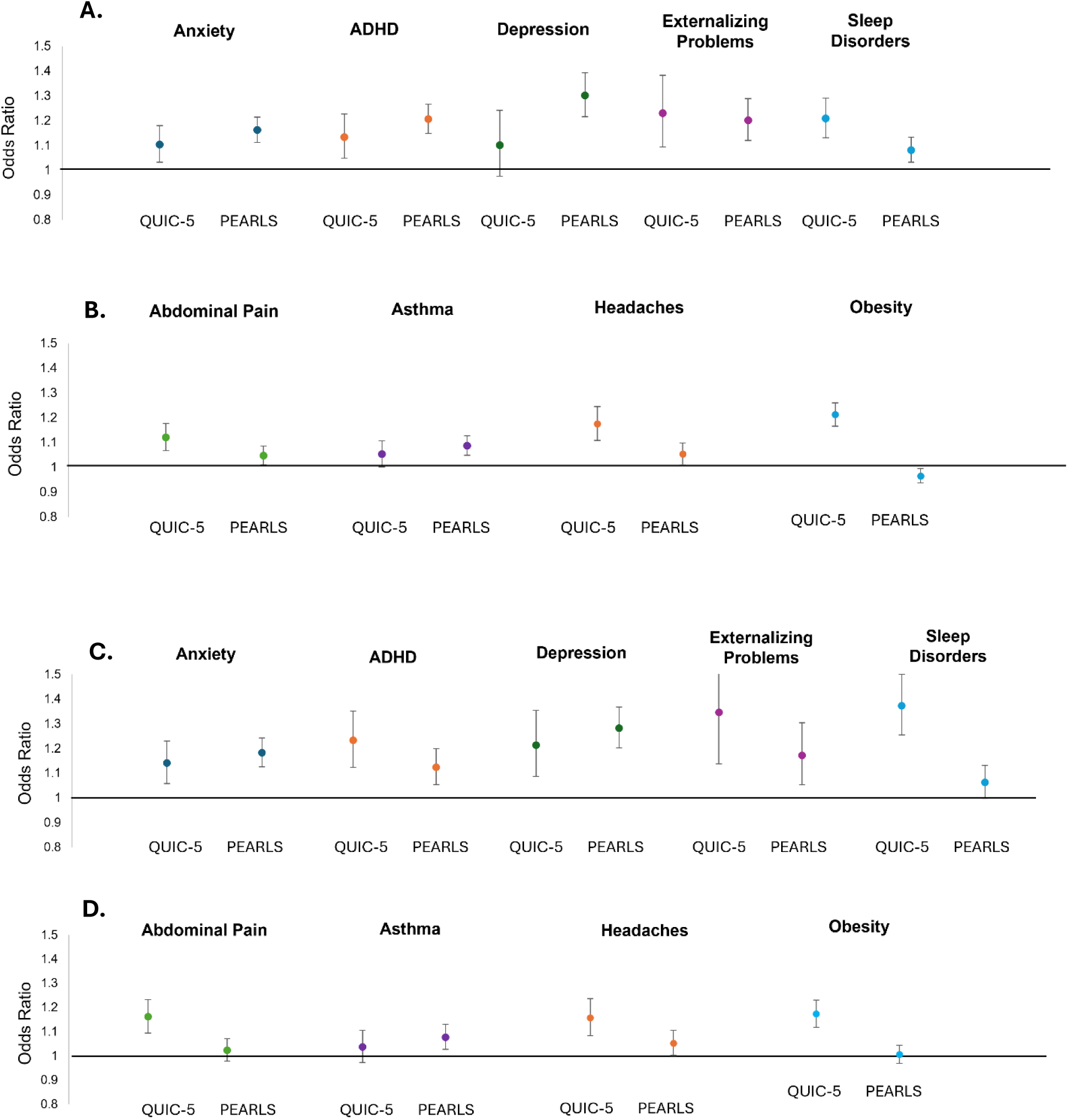
ACEs (PEARLS) and unpredictability (QUIC-5) screens independently predict child mental and physical health outcomes. Odds ratios and 95% confidence intervals. Panels A-B show associations for caregiver reports. Youth self-report in panels C-D.

The QUIC identified significant risk for health problems that is not captured by the PEARLS screen alone also when a second analytic approach was used (Figure 6). For example, examining risk for depression, Figure 6 shows the odds ratios associated with all possible combinations of QUIC and PEARLS scores. Children with a score of 4 or more on the PEARLS had an increased risk of depression. For example, the odds ratio for a PEARLS score of 4 or more and a QUIC score of zero was 7.9. However, a child with a score of zero on the PEARLS and 4 on the QUIC is 11.8 more likely to have a depression diagnosis than a child who scores a zero on both screens. This indicates that the QUIC-5 identifies a significant population of children with mental health vulnerabilities derived from unpredictable environments that would be missed by restricting screening to current ACEs tools.

**Figure 6.**
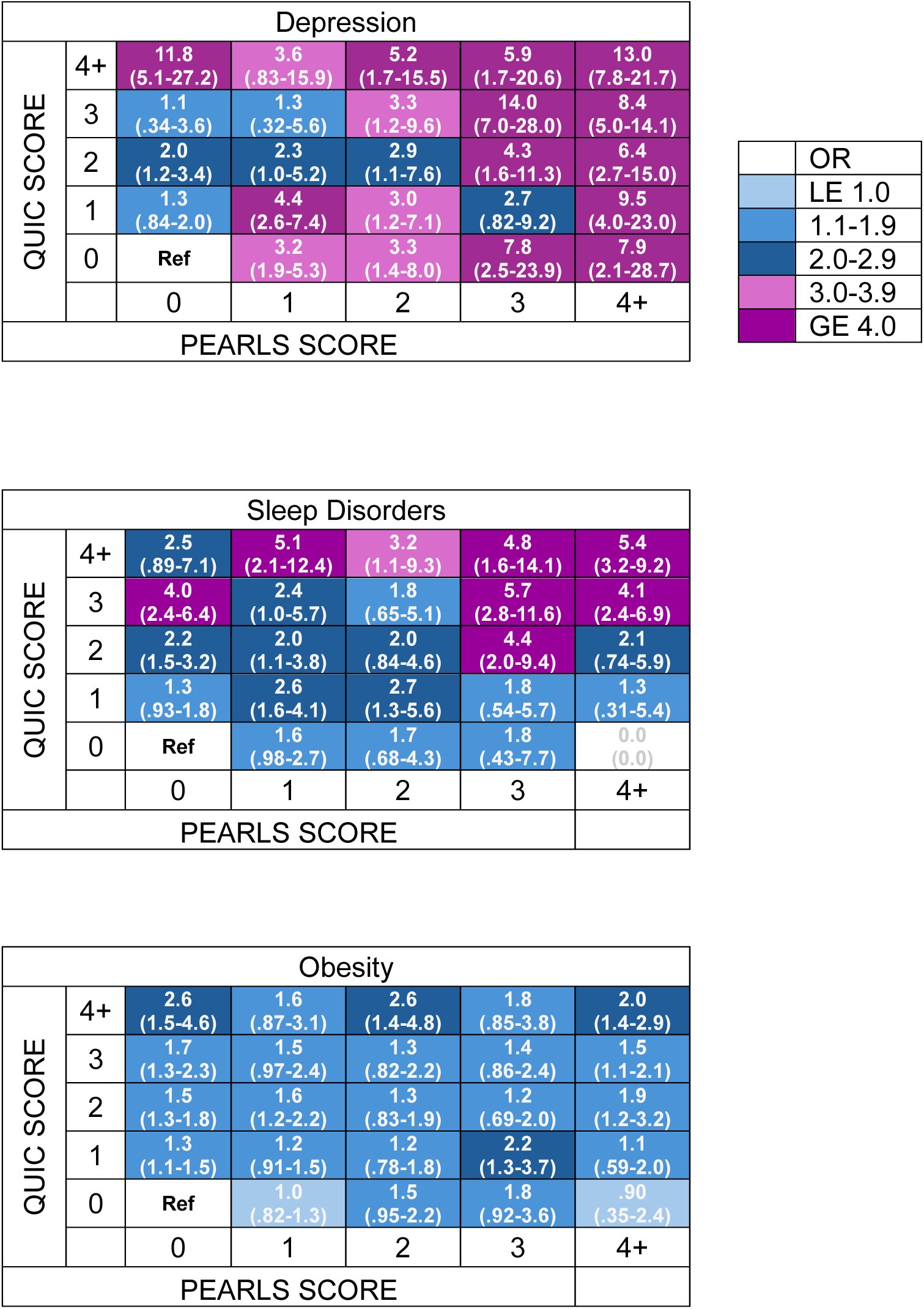
Evaluation of the independent and combined predictive power of ACEs (PEARLS) and unpredictability (QUIC) screens. Examples shown here are for youth self-report. Results for both youth and caregiver reports for all diagnoses can be found in Table S11.

This observation was not unique to depression. For example, a child with a “high score” on the PEARLS (4 or more) and with a score of 1 on the QUIC, is not at increased risk of a sleep disorder. However, a child who meets this criterion on the PEARLS and has a QUIC score of 4 or more is 5.4 times more likely to have a sleep disorder diagnosis than a child with a zero score on both screens (similar heatmaps for all diagnoses can be found in Table S11).

## Discussion

The principal findings of this study conducted in more than 29,000 children at 19 pediatric primary care clinics are: (a) Widespread, systematic screening for ACEs and early-life unpredictability is feasible in primary care clinics serving diverse communities. (b) Exposure to adversity measured with both the QUIC and PEARLS is prevalent even among the youngest children and increases with age. (c) Although parent and youth reports are in general agreement and both predict health risks, youth report more exposures. (d) The unpredictability screen (QUIC-5) portends child outcomes at least as well, and in some cases better, than currently recommended ACEs screens. (e) Crucially, for some health problems (e.g. depression, obesity), the QUIC-5 identifies risk for mental and physical health problems that is not captured by the PEARLS screen.

Our study was conducted in Orange County, California (CA) which is home to the third largest child population in the state (after Los Angeles and San Diego counties). The youth served by CHOC are representative of the diverse socioeconomic, ethnic and cultural constituency of both the county and of California more broadly. Underscoring the representative nature of the sample is the fact that our race and ethnic distributions largely mirror that of both the county and state^28^. This indicates that both the feasibility and uptake of the screening as well as the associations we observed with child health outcomes are highly likely to generalize.

A concerning observation was the prevalence of adversity exposures even among the youngest children, with 13 percent already having experienced at least one ACE and 27 percent exposed to at least one form of unpredictability before 5 years of age. As children aged, both caregivers and youth reported more exposures. On average, youths reported more exposures to both ACEs and unpredictability than did caregivers, highlighting the importance of obtaining youth self-report in addition to caregiver report when possible. According to self-reports, by late adolescence 27 percent of youth had been exposed to one or more ACE and 49 percent at least one form of unpredictability.

For both mental and physical health outcomes, exposure to more ACEs and unpredictability exhibited a graded relationship with risk of diagnosis. Additionally, the predictive power of unpredictability was similar in magnitude to ACEs for the range of mental and physical health outcomes examined. The results strongly suggest that consideration of unpredictability enhances the ability to identify children at risk of a mental or physical health condition beyond that of the PEARLS: First, when modeling the two screens together, each predicted risk independently from the other for the mental and physical health outcomes examined. Furthermore, high QUIC scores predicted significant risk for many diagnoses even when the PEARLS score was low. These risks (in some cases, odds ratios greater than 10) would be missed if PEARLS scores alone were used for screening.

For two health conditions, obesity and sleep disorders, the QUIC was superior in risk identification compared to the PEARLS. This observation regarding obesity mirrors findings from a study of 367 children receiving care at a safety net practice in Northern California that documented positive associations between caregiver reports with the PEARLS and asthma, but not obesity^29^. This is not entirely surprising as healthy eating and sleep hygiene are both dependent on structure and routines^30,31^. Delineating both the shared and unique contributions of different forms of early life adversity to individual disease burden, as done here, paves the path for more precise understanding of individual exposures, and the mechanisms through which those exposures operate, enabling true precision medicine.

An additional strength of the current study was the examination of the utility of youth self-report of exposure to adversity. Following State of California recommendations for the PEARLS, children aged 12 and above completed both screens. In general, the dose-response patterns associated with both mental and physical health outcomes were comparable for youth and caregiver reports, including the findings that the QUIC was a better predictor of sleep disorders and obesity than the PEARLS. These similarities underscore the validity of youth report on the QUIC and PEARLS and increase confidence in the validity of both screening instruments. A limitation of the study lies in the reliance on child health diagnoses derived from electronic health records. Given the study design, it is not possible to disentangle the temporal relations between exposures and diagnoses. Further, particularly for the mental health outcomes, we are unable to probe the associations between exposures and subclinical symptom profiles, which are highly likely. Future studies will explore how structural and social determinants of health influence exposures to unpredictability and their associations with health, to better identify inequities which exist. This is of particular importance because structural determinants of ACEs and unpredictability are not evenly distributed and individuals of historically and currently marginalized and systematically excluded backgrounds are at disproportionate risk for these exposures^32–36^.

There are several features of unpredictability that distinguish it from many other forms of early life adversity, rendering it a promising target for prevention and intervention: (1) There are multiple opportunities for prevention and intervention, including the prospect of multilevel intervention^37,38^. These range from the level of the individual (e.g. altering caregiver attitudes regarding the importance of predictability), to the family systems level (e.g. implementing family routines) and at the public policy level (e.g. adoption of Fair Workweek regulations that address precarious parental work schedules). Addressing the full range of ACEs such as poverty and abuse should be a top priority to improve the well-being of children and families. However, a parallel and attainable goal is increasing predictability in children’s lives, for example through the encouragement of family routines, which have the additional advantage of being relatively low cost. (2) In conceptualizing high unpredictability as a form of early life adversity, it is notable that low unpredictability (i.e., predictable environments) may exert protective influences on child development and buffer children from adversity. For example, in families experiencing poverty, parental substance use disorders, chronic illness or divorce, family routines predict child resilience^39–41^. In addition, predictability protected child mental health in the context of societal-level disruptions such as the COVID-19 pandemic^42,43^. (3) Community perspectives obtained from pediatricians, allied health care providers and parents indicate that screening in pediatric primary care with the QUIC-5 may be more acceptable than screening with the PEARLS^44^. (4) Finally, interventions seeking to promote predictability should and can incorporate culturally-responsive, person-centered, and community inclusive approaches with the acknowledgment that the promotion of predictability requires consideration of the unique needs of each child and family.

## Conclusion

In sum, we demonstrate that large-scale systematic screening for ACES and early-life unpredictability is feasible in primary care clinics serving diverse communities. Exposures to these types of adversity are prevalent even among the youngest children, and both predict health outcomes. The unpredictability screen (QUIC-5) portends child outcomes at least as well, and in some cases better, than current ACEs screening (PEARLS-tool) and for some health problems, the QUIC-5 identifies risk for mental and physical health problems that are not captured by the ACEs screen These findings suggest that consideration of unpredictability will (1) identify children at risk for health problems that will be otherwise missed and (2) may provide a tractable target for prevention and intervention in the pediatric primary care setting with the potential to make meaningful positive impacts on child and life-span health.

## Data Availability

All data produced in the present study are available upon reasonable request to the authors.

## ACKNOWLEDGEMENTS

The authors are grateful to the families who participated, to the valuable guidance provided by the SoCal Kids Study Community Advisory Board, and to the dedicated staff at the Early Human and Lifespan Development Program and Children’s Hospital of Orange County whose efforts made this work possible.

