## Supplemental Materials for "Contribution of an under-recognized adversity to child health risk: large-scale, population-based ACEs screening"

Table S1. ICD-10 codes used to define mental and physical health diagnoses.

| Mental Health | | Physical Health | |
| --- | --- | --- | --- |
| ADHD | F90.0-F90.2, F90.8, F90.9 | **Abdominal Pain** | R10 |
| Anxiety | F40.00-F40.02, F40.10, F40.11, F40.210, F40.218, F40.220, F40.228, F40.230-F40.233, F40.240-F40.243, F40.248, F40.290, F40.291, F40.298, F40.8, F40.9, F41.0-F41.3, F41.8, F41.9, F93.0, F94.0 | **Asthma** | J45 |
| Depression | F32.0-F32.5, F32.81, F32.89, F32.9, F33.0-F33.3, F33.40-F33.42, F33.8, F33.9, F34.1, F348, F34.81 | **Headache/Migraine** | G43, G44, R51 |
| Externalizing Problems | F63.0-F63.2, F63.81, F63.89, F63.9, F91.0-F91.3, F91.8, F91.9 | **Obesity** | E66, Z68.53, Z68.54 |
| Sleep Disorders | G47 |  |  |

Table S2. Incidence of child mental and physical heath diagnoses.

| Diagnosis | n | % |
| --- | --- | --- |
| Abdominal Pain |  |  |
| yes | 4,143 | 14.1 |
| no | 25,612 | 85.9 |
| Anxiety |  |  |
| yes | 1,890 | 6.4 |
| no | 27,495 | 96.3 |
| Asthma |  |  |
| yes | 4,705 | 13.9 |
| no | 25,230 | 86.1 |
| ADHD |  |  |
| yes | 1,232 | 4.2 |
| no | 28,073 | 95.8 |
| Depression |  |  |
| yes | 545 | 1.9 |
| no | 28,760 | 98.1 |
| Externalizing Problems |  |  |
| yes | 494 | 1.7 |
| no | 28,811 | 98.3 |
| Headache |  |  |
| yes | 2,562 | 8.7 |
| no | 26,743 | 91.3 |
| Obesity |  |  |
| yes | 9,402 | 32.1 |
| no | 19,903 | 67.9 |
| Sleep Disorders |  |  |
| yes | 1,812 | 6.2 |
| no | 27,493 | 93.8 |

Table S3. Caregiver endorsement for exposures on PEARLS and QUIC-5 by child age group.

|  | Mean | SD |
| --- | --- | --- |
| PEARLS |  |  |
| 0 to 4 | .24 | .77 |
| 5 to 9 | .38 | 1.1 |
| 10-14 | .47 | 1.2 |
| 15-18 | .54 | 1.3 |
| QUIC-5 |  |  |
| 0 to 4 | .34 | .72 |
| 5 to 9 | .43 | .82 |
| 10-14 | .60 | 1.4 |
| 15-18 | .67 | .98 |

Table S4. Youth and caregiver endorsement on PEARLS and QUIC for youth ages 12-18. These are for the subset of caregivers and youth pairs both providing data (n = 8,629 for PEARLS and n= 7,369 for QUIC).

|  | Mean | SD |
| --- | --- | --- |
| PEARLS |  |  |
| Youth | .6 | 1.4 |
| Caregiver | .51 | 1.3 |
| QUIC-5 |  |  |
| Youth | .86 | 1.1 |
| Caregiver | .60 | .94 |

| Table S5. Caregiver report on PEARLS (n = 20,261) and QUIC (n = 26,804) screeners predict child mental health conditions. | | | | | | | | | |
| --- | --- | --- | --- | --- | --- | --- | --- | --- | --- |
|  | **PEARLS** | | | |  | **QUIC-5** | | | |
| Diagnosis | Score | Adjusted Odds Ratio | 95% Confidence Interval | p-value |  | Score | Adjusted Odds Ratio | 95% confidence Interval | p-value |
| Anxiety | 0 | 1.0 | Referent |  |  | 0 | 1.0 | Referent |  |
|  | 1 | 1.6 | (1.3-1.9) | .00 |  | 1 | 1.2 | (1.1-1.4) | .01 |
|  | 2 | 2.1 | (1.7-2.7) | .00 |  | 2 | 1.5 | (1.2-1.7) | .00 |
|  | 3 | 1.6 | (1.2-2.3) | .00 |  | 3 | 1.9 | (1.6-2.4) | .00 |
|  | 4 or more | 3.1 | (2.5-3.8) | .00 |  | 4 or more | 1.7 | (1.2-2.6) | .01 |
| ADHD | 0 | 1.0 | Referent |  |  | 0 | 1.0 | Referent |  |
|  | 1 | 1.7 | (1.4-2.1) | .00 |  | 1 | 1.3 | (1.1-1.5) | .00 |
|  | 2 | 2.3 | (1.8-3.1) | .00 |  | 2 | 1.5 | (1.3-1.9) | .00 |
|  | 3 | 2.3 | (1.6-3.3) | .00 |  | 3 | 2.7 | (2.2-3.4) | .00 |
|  | 4 or more | 3.7 | (2.9-4.8) | .00 |  | 4 or more | 3.1 | (2.1-4.6) | .00 |
| Depression | 0 | 1.0 | Referent |  |  | 0 | 1.0 | Referent |  |
|  | 1 | 2.1 | (1.6-2.9) | .00 |  | 1 | 1.5 | (1.2-1.9) | .00 |
|  | 2 | 3.3 | (2.3-4.8) | .00 |  | 2 | 2.3 | (1.7-3.0) | .00 |
|  | 3 | 3.4 | (2.1-5.5) | .00 |  | 3 | 2.5 | (1.7-3.5) | .00 |
|  | 4 or more | 5.4 | (3.9-7.4) | .00 |  | 4 or more | 3.2 | (1.8-5.7) | .00 |
| Externalizing  Problems | 0 | 1.0 | Referent |  |  | 0 | 1.0 | Referent |  |
|  | 1 | 2.2 | (1.6-2.9) | .00 |  | 1 | 1.4 | (1.1-1.7) | .01 |
|  | 2 | 1.8 | (1.1-2.9) | .02 |  | 2 | 2.0 | (1.5-2.7) | .00 |
|  | 3 | 2.7 | (1.6-4.6) | .00 |  | 3 | 3.1 | (2.2-4.3) | .00 |
|  | 4 or more | 4.5 | (3.1-6.5) | .00 |  | 4 or more | 3.6 | (2.0-6.5) | .00 |
| Sleep Disorders | 0 | 1.0 | Referent |  |  | 0 | 1.0 | Referent |  |
|  | 1 | 1.4 | (1.2-1.7) | .00 |  | 1 | 1.3 | (1.1-1.4) | .00 |
|  | 2 | 1.7 | (1.3-2.2) | .00 |  | 2 | 1.7 | (1.4-1.9) | .00 |
|  | 3 | 2.1 | (1.5-2.8) | .00 |  | 3 | 2.1 | (1.7-2.6) | .00 |
|  | 4 or more | 1.9 | (1.5-2.5) | .00 |  | 4 or more | 2.5 | (1.7-3.6) | .00 |
| Note: Odds ratio estimates adjusted for child age, child age* and sex. | | | | | | | | | |

| Table S6. Youth self-report on PEARLS (n = 8,906) and QUIC (n = 9,870) screeners predict youth mental health conditions. | | | | | | | | | |
| --- | --- | --- | --- | --- | --- | --- | --- | --- | --- |
|  | **PEARLS** | | | |  | **QUIC-5** | | | |
| Diagnosis | Score | Adjusted Odds Ratio | 95% Confidence Interval | p-value |  | Score | Adjusted Odds Ratio | 95% confidence Interval | p-value |
| Anxiety | 0 | 1.0 | Referent |  |  | 0 | 1.0 | Referent |  |
|  | 1 | 1.7 | (1.4-2.0) | .00 |  | 1 | 1.2 | (.99-1.4) | .07 |
|  | 2 | 1.9 | (1.5-2.5) | .00 |  | 2 | 1.6 | (1.3-1.9) | .00 |
|  | 3 | 3.0 | (2.3-4.1) | .00 |  | 3 | 2.0 | (1.6-2.5) | .00 |
|  | 4 or more | 3.7 | (2.9-4.7) | .00 |  | 4 or more | 3.3 | (2.5-4.3) | .00 |
| ADHD | 0 | 1.0 | Referent |  |  | 0 | 1.0 | Referent |  |
|  | 1 | 1.5 | (1.1-1.9) | .01 |  | 1 | 1.4 | (1.1-1.7) | .00 |
|  | 2 | 2.2 | (1.5-3.1) | .00 |  | 2 | 1.9 | (1.4-2.4) | .00 |
|  | 3 | 2.2 | (1.4-3.4) | .00 |  | 3 | 2.1 | (1.6-2.9) | .00 |
|  | 4 or more | 3.2 | (2.3-4.4) | .00 |  | 4 or more | 3.0 | (2.1-4.4) | .00 |
| Depression | 0 | 1.0 | Referent |  |  | 0 | 1.0 | Referent |  |
|  | 1 | 2.7 | (2.0-3.6) | .00 |  | 1 | 1.4 | (1.1-1.9) | .01 |
|  | 2 | 2.8 | (1.9-4.2) | .00 |  | 2 | 2.1 | (1.6-2.9) | .00 |
|  | 3 | 5.1 | (3.4-7.5) | .00 |  | 3 | 3.7 | (2.7-5.0) | .00 |
|  | 4 or more | 7.2 | (5.3-9.7) | .00 |  | 4 or more | 6.1 | (4.3-8.7) | .00 |
| Externalizing Problems | 0 | 1.0 | Referent |  |  | 0 | 1.0 | Referent |  |
|  | 1 | 2.3 | (1.4-3.6) | .00 |  | 1 | 1.7 | (1.1-2.5) | .02 |
|  | 2 | 2.7 | (1.5-5.1) | .00 |  | 2 | 2.7 | (1.7-4.3) | .00 |
|  | 3 | 4.7 | (2.5-8.8) | .00 |  | 3 | 3.3 | (2.0-5.6) | .00 |
|  | 4 or more | 4.9 | (2.9-8.4) | .00 |  | 4 or more | 4.7 | (2.5-8.7) | .00 |
| Sleep Disorders | 0 | 1.0 | Referent |  |  | 0 | 1.0 | Referent |  |
|  | 1 | 1.6 | (1.2-2.1) | .00 |  | 1 | 1.3 | (1.0-1.6) | .03 |
|  | 2 | 1.6 | (1.1-2.3) | .01 |  | 2 | 1.9 | (1.5-2.4) | .00 |
|  | 3 | 2.7 | (1.9-4.0) | .00 |  | 3 | 2.6 | (2.0-3.5) | .00 |
|  | 4 or more | 2.6 | (1.9-3.6) | .00 |  | 4 or more | 3.0 | (2.2-4.3) | .00 |
| Note: Odds ratio estimates adjusted for child age and sex. | | | | | | | | | |

| Table S7. Caregiver report on PEARLS (n = 20,261) and QUIC (n = 26,804) screeners predict child physical health conditions. | | | | | | | | | |
| --- | --- | --- | --- | --- | --- | --- | --- | --- | --- |
|  | **PEARLS** | | | |  | **QUIC-5** | | | |
| Diagnosis | Score | Adjusted Odds Ratio | 95% Confidence Interval | p-value |  | Score | Adjusted Odds Ratio | 95% confidence Interval | p-value |
| Abdominal Pain | 0 | 1.0 | Referent |  |  | 0 | 1.0 | Referent |  |
|  | 1 | 1.2 | (1.0-1.3) | .01 |  | 1 | 1.4 | (1.3-1.5) | .00 |
|  | 2 | 1.3 | (1.1-1.6) | .00 |  | 2 | 1.4 | (1.3-1.6) | .00 |
|  | 3 | 1.4 | (1.1-1.7) | .02 |  | 3 | 1.5 | (1.3-1.8) | .00 |
|  | 4 or more | 1.6 | (1.3-2.0) | .00 |  | 4 or more | 1.6 | (1.1-2.1) | .01 |
| Asthma | 0 | 1.0 | Referent |  |  | 0 | 1.0 | Referent |  |
|  | 1 | 1.1 | (1.0-1.3) | .06 |  | 1 | 1.1 | (1.0-1.2) | .16 |
|  | 2 | 1.5 | (1.2-1.8) | .00 |  | 2 | 1.3 | (1.1-1.4) | .00 |
|  | 3 | 1.6 | (1.2-2.0) | .00 |  | 3 | 1.4 | (1.2-1.7) | .00 |
|  | 4 or more | 1.6 | (1.3-2.0) | .00 |  | 4 or more | 1.6 | (1.2-2.2) | .00 |
| Headache | 0 | 1.0 | Referent |  |  | 0 | 1.0 | Referent |  |
|  | 1 | 1.3 | (1.2-1.6) | .00 |  | 1 | 1.2 | (1.1-1.4) | .00 |
|  | 2 | 1.5 | (1.2-1.9) | .00 |  | 2 | 1.5 | (1.3-1.7) | .00 |
|  | 3 | 1.2 | (.87-1.6) | .27 |  | 3 | 2.0 | (1.7-2.4) | .00 |
|  | 4 or more | 1.9 | (1.5-2.3) | .00 |  | 4 or more | 1.9 | (1.4-2.7) | .00 |
| Obesity |  |  |  |  |  |  |  |  |  |
|  | 0 | 1.0 | Referent |  |  | 0 | 1.0 | Referent |  |
|  | 1 | .99 | (.89-1.1) | .80 |  | 1 | 1.3 | (1.2-1.4) | .00 |
|  | 2 | 1.1 | (.91-1.2) | .42 |  | 2 | 1.5 | (1.4-1.6) | .00 |
|  | 3 | 1.0 | (.82-1.2) | .95 |  | 3 | 1.6 | (1.4-1.8) | .00 |
|  | 4 or more | 1.4 | (1.2-1.6) | .00 |  | 4 or more | 1.7 | (1.3-2.2) | .00 |
| Note: Odds ratio estimates adjusted for child age, child age* and sex. | | | | | | | | | |

| Table S8. Youth self-report on PEARLS (n = 8,906) and QUIC (n = 9,870) screeners predict child physical health conditions. | | | | | | | | | |
| --- | --- | --- | --- | --- | --- | --- | --- | --- | --- |
|  | **PEARLS** | | | |  | **QUIC-5** | | | |
| Diagnosis | Score | Adjusted Odds Ratio | 95% Confidence Interval | p-value |  | Score | Adjusted Odds Ratio | 95% confidence Interval | p-value |
| Abdominal Pain | 0 | 1.0 | Referent |  |  | 0 | 1.0 | Referent |  |
|  | 1 | 1.4 | (1.2-1.6) | .01 |  | 1 | 1.4 | (.96-1.0) | .00 |
|  | 2 | 1.4 | (1.2-1.8) | .00 |  | 2 | 1.6 | (1.4-1.9) | .00 |
|  | 3 | 1.5 | (1.1-2.0) | .14 |  | 3 | 1.6 | (1.3-1.9) | .00 |
|  | 4 or more | 1.7 | (1.3-2.1) | .00 |  | 4 or more | 2.0 | (1.6-2.6) | .00 |
| Asthma | 0 | 1.0 | Referent |  |  | 0 | 1.0 | Referent |  |
|  | 1 | 1.1 | (.90-1.3) | .52 |  | 1 | 1.1 | (.95-1.2) | .25 |
|  | 2 | 1.5 | (1.2-1.9) | .01 |  | 2 | 1.3 | (1.1-1.5) | .00 |
|  | 3 | 1.4 | (1.0-1.9) | .40 |  | 3 | 1.3 | (1.0-1.6) | .03 |
|  | 4 or more | 1.4 | (1.1-1.8) | .00 |  | 4 or more | 1.5 | (1.2-2.0) | .00 |
| Headache | 0 | 1.0 | Referent |  |  | 0 | 1.0 | Referent |  |
|  | 1 | 1.5 | (1.2-1.8) | .00 |  | 1 | 1.2 | (1.2-1.6) | .00 |
|  | 2 | 1.4 | (1.1-1.8) | .01 |  | 2 | 1.3 | (1.3-1.8) | .00 |
|  | 3 | 1.6 | (1.1-2.1) | .01 |  | 3 | 1.4 | (1.4-2.2) | .00 |
|  | 4 or more | 1.7 | (1.4-2.3) | .00 |  | 4 or more | 1.6 | (1.6-2.7) | .00 |
| Obesity | 0 | 1.0 | Referent |  |  | 0 | 1.0 | Referent |  |
|  | 1 | 1.1 | (.98-1.3) | .23 |  | 1 | 1.3 | (1.2-1.5) | .00 |
|  | 2 | 1.3 | (1.0-1.5) | .36 |  | 2 | 1.5 | (1.4-1.7) | .00 |
|  | 3 | 1.3 | (1.1-1.7) | .12 |  | 3 | 1.6 | (1.3-1.9) | .00 |
|  | 4 or more | 1.4 | (1.2-1.7) | .00 |  | 4 or more | 2.1 | (1.7-2.6) | .00 |
| Note: Odds ratio estimates adjusted for child age and sex. | | | | | | | | | |

| Table S9. Binary logistic regressions with PEARLS and QUIC caregiver scores (continuous) concurrently entered as predictors (n = 18,486). | | | | |
| --- | --- | --- | --- | --- |
| Diagnosis | Score | Adjusted Odds Ratio | 95% Confidence Interval | p-value |
| Abdominal Pain |  |  |  |  |
|  | PEARLS | 1.0 | (1.0-1.1) | .01 |
|  | QUIC | 1.1 | (1.1-1.2) | .00 |
| Anxiety |  |  |  |  |
|  | PEARLS | 1.2 | (1.1-1.2) | .00 |
|  | QUIC | 1.1 | (1.0-1.2) | .00 |
| Asthma |  |  |  |  |
|  | PEARLS | 1.1 | (1.0-1.1) | .00 |
|  | QUIC | 1.1 | (1.0-1.1) | .04 |
| ADHD |  |  |  |  |
|  | PEARLS | 1.2 | (1.1-1.3) | .00 |
|  | QUIC | 1.1 | (1.0-1.2) | .01 |
| Depression |  |  |  |  |
|  | PEARLS | 1.3 | (1.2-1.4) | .00 |
|  | QUIC | 1.1 | (.98-1.2) | .12 |
| Externalizing Problems |  |  |  |  |
|  | PEARLS | 1.2 | (1.1-1.3) | .00 |
|  | QUIC | 1.2 | (1.1-1.4) | .00 |
| Headache |  |  |  |  |
|  | PEARLS | 1.1 | (1.0-1.1) | .02 |
|  | QUIC | 1.2 | (1.1-1.2) | .00 |
| Obesity |  |  |  |  |
|  | PEARLS | .97 | (.94-1.0) | .03 |
|  | QUIC | 1.2 | (1.2-1.3) | .00 |
| Sleep Disorders |  |  |  |  |
|  | PEARLS | 1.1 | (1.0-1.1) | .00 |
|  | QUIC | 1.2 | (1.1-1.3) | .00 |

| Table S10. Binary logistic regressions with PEARLS and QUIC youth scores (continuous) concurrently entered as predictors (n = 7,149). | | | | |
| --- | --- | --- | --- | --- |
| Diagnosis | Score | Adjusted Odds Ratio | 95% Confidence Interval | p-value |
| Abdominal Pain |  |  |  |  |
|  | PEARLS | 1.0 | (.98-1.1) | .32 |
|  | QUIC | 1.2 | (1.1-1.2) | .00 |
| Anxiety |  |  |  |  |
|  | PEARLS | 1.2 | (1.1-1.2) | .00 |
|  | QUIC | 1.1 | (1.1-1.2) | .00 |
| Asthma |  |  |  |  |
|  | PEARLS | 1.1 | (1.0-1.1) | .00 |
|  | QUIC | 1.0 | (.98-1.1) | .22 |
| ADHD |  |  |  |  |
|  | PEARLS | 1.1 | (1.1-1.2) | .00 |
|  | QUIC | 1.2 | (1.1-1.4) | .00 |
| Depression |  |  |  |  |
|  | PEARLS | 1.3 | (1.2-1.4) | .00 |
|  | QUIC | 1.2 | (1.1-1.4) | .00 |
| Externalizing Problems |  |  |  |  |
|  | PEARLS | 1.2 | (1.1-1.3) | .00 |
|  | QUIC | 1.3 | (1.1-1.6) | .00 |
| Headache |  |  |  |  |
|  | PEARLS | 1.1 | (1.0-1.1) | .04 |
|  | QUIC | 1.2 | (1.1-1.2) | .00 |
| Obesity |  |  |  |  |
|  | PEARLS | 1.0 | (.97-1.0) | .76 |
|  | QUIC | 1.2 | (1.1-1.2) | .00 |
| Sleep Disorders |  |  |  |  |
|  | PEARLS | 1.1 | (1.0-1.1) | .06 |
|  | QUIC | 1.4 | (1.3-1.5) | .00 |

Table S11. Data summary of the combined and unique predictive contributions of the PEARLS and QUIC-5.

| Caregiver Reports | | | | | | |
| --- | --- | --- | --- | --- | --- | --- |
| Anxiety | | | | | | |
| QUIC | 4+ | **1.6**  **(.63-4.2)** | **0.0**  **(0.0)** | **1.8**  **(.42-8.1)** | **2.0**  **(0.6-6.8)** | **3.1**  **(1.7-5.6)** |
|  | 3 | **1.8**  **(1.2-2.7)** | **1.9**  **(1.0-3.7)** | **5.8**  **(3.1-10.6)** | **1.3**  **(0.5-3.6)** | **2.4**  **(1.6-3.6)** |
|  | 2 | **1.5**  **(1.1-1.9)** | **1.7**  **(1.0-2.8)** | **1.4**  **(0.7-2.7)** | **2.2**  **(1.1-4.4)** | **4.4**  **(2.7-7.3)** |
|  | 1 | **1.2**  **(1.0-1.4)** | **2.0**  **(1.5-2.8)** | **2.2**  **(1.4-3.4)** | **1.3**  **(0.6-2.7)** | **4.0**  **(2.3-6.9)** |
|  | 0 | **Ref** | **1.6**  **(1.2-2.0)** | **2.0**  **(1.3-3.0)** | **2.2**  **(1.1-4.2)** | **3.4**  **(1.9-6.1)** |
|  |  | 0 | 1 | 2 | 3 | 4+ |
| PEARLS | | | | | |  |

| Youth Self-Reports | | | | | | |
| --- | --- | --- | --- | --- | --- | --- |
| Anxiety | | | | | | |
| QUIC | 4+ | **1.7**  **(.67-4.4)** | **2.4**  **(.92-6.2)** | **2.9**  **(1.3-6.7)** | **1.7**  **(.50-5.6)** | **6.3**  **(4.2-9.5)** |
|  | 3 | **1.8**  **(1.1-3.0)** | **1.7**  **(.82-3.7)** | **2.2**  **(1.1-4.5)** | **3.7**  **(2.0-7.1)** | **3.6**  **(2.4-5.5)** |
|  | 2 | **1.8**  **(1.3-2.4)** | **1.9**  **(1.2-3.2)** | **1.3**  **(.64-2.8)** | **2.7**  **(1.4-5.5)** | **4.8**  **(2.7-8.7)** |
|  | 1 | **1.3**  **(1.0-1.7)** | **1.4**  **(.92-2.2)** | **2.1**  **(1.1-3.8)** | **4.0**  **(2.1-7.7)** | **2.6**  **(1.1-5.9)** |
|  | 0 | **Ref** | **2.2**  **(1.5-3.1)** | **2.6**  **(1.4-4.6)** | **5.6**  **(2.5-12.3)** | **1.4**  **(.33-6.3)** |
|  |  | 0 | 1 | 2 | 3 | 4+ |
| PEARLS | | | | | |  |

| ADHD | | | | | | |
| --- | --- | --- | --- | --- | --- | --- |
| QUIC | 4+ | **2.4**  **(.83-6.8)** | **6.0**  **(2.6-14.2)** | **4.5**  **(1.7-11.8)** | **1.1**  **(.15-8.2)** | **4.5**  **(2.4-8.2)** |
|  | 3 | **2.5**  **(1.4-4.3)** | **1.8**  **(.70-4.5)** | **2.5**  **(.97-6.4)** | **3.5**  **(1.5-8.5)** | **4.3**  **(2.5-7.6)** |
|  | 2 | **2.3**  **(1.6-3.2)** | **1.7**  **(.90-3.3)** | **2.3**  **(1.0-5.1)** | **.99**  **(.24-4.1)** | **5.0**  **(2.2-11.5)** |
|  | 1 | **1.5**  **(1.1-2.0)** | **2.3**  **(1.4-3.8)** | **3.9**  **(1.9-7.8)** | **3.4**  **(1.3-8.8)** | **2.0**  **(.59-6.5)** |
|  | 0 | **Ref** | **1.3**  **(.77-2.3)** | **3.0**  **(1.4-6.4)** | **5.3**  **(2.0-14.1)** | **6.2**  **(1.8-22.1)** |
|  |  | 0 | 1 | 2 | 3 | 4+ |
| PEARLS | | | | | |  |

| ADHD | | | | | | |
| --- | --- | --- | --- | --- | --- | --- |
| QUIC | 4+ | **3.1**  **(1.2-7.9)** | **0.0**  **(0.0)** | **2.8**  **(.62-12.3)** | **4.5**  **(1.5-13.5)** | **4.3**  **(2.2-8.3)** |
|  | 3 | **2.1**  **(1.3-3.4)** | **2.2**  **(.99-4.8)** | **2.6**  **(1.0-6.5)** | **1.4**  **(.44-4.7)** | **3.3**  **(2.1-5.2)** |
|  | 2 | **1.2**  **(.85-1.6)** | **2.8**  **(1.7-4.6)** | **2.0**  **(.96-4.2)** | **1.5**  **(.53-4.1)** | **3.1**  **(1.6-6.0)** |
|  | 1 | **1.2**  **(1.0-1.5)** | **2.1**  **(1.4-3.0)** | **2.5**  **(1.5-4.1)** | **2.7**  **(1.3-5.4)** | **3.7**  **(1.9-7.2)** |
|  | 0 | **Ref** | **1.4**  **(1.0-1.9)** | **2.8**  **(1.8-4.4)** | **2.2**  **(1.1-4.7)** | **4.6**  **(2.4-8.8)** |
|  |  | 0 | 1 | 2 | 3 | 4+ |
| PEARLS | | | | | |  |

|  | OR |
| --- | --- |
|  | LE 1.0 |
|  | 1.1-1.9 |
|  | 2.0-2.9 |
|  | 3.0-3.9 |
|  | GE 4.0 |

| Depression | | | | | | |
| --- | --- | --- | --- | --- | --- | --- |
| QUIC | 4+ | **1.2**  **(.16-9.3)** | **4.4**  **(.55-34.7)** | **0.0**  **(0.0)** | **7.5**  **(1.6-34.8)** | **6.0**  **(2.6-13.9)** |
|  | 3 | **2.0**  **(.88-4.3)** | **1.8**  **(.55-6.0)** | **4.9**  **(1.4-16.9)** | **3.7**  **(.86-16.1)** | **5.4**  **(3.0-9.6)** |
|  | 2 | **2.1**  **(1.3-3.3)** | **.92**  **(.22-3.8)** | **2.3**  **(.80-6.4)** | **10.2**  **(4.4-23.2)** | **7.6**  **(3.5-16.3)** |
|  | 1 | **1.2**  **(.80-1.8)** | **2.8**  **(1.6-5.0)** | **5.4**  **(3.0-10.0)** | **2.2**  **(.66-7.1)** | **6.8**  **(3.0-15.3)** |
|  | 0 | **Ref** | **2.5**  **(1.6-4.0)** | **3.3**  **(1.6-6.8)** | **2.9**  **(.88-9.7)** | **4.1**  **(1.4-11.9)** |
|  |  | 0 | 1 | 2 | 3 | 4+ |
| PEARLS | | | | | |  |

| Depression | | | | | | |
| --- | --- | --- | --- | --- | --- | --- |
| QUIC | 4+ | **11.7**  **(5.1-27.2)** | **3.6**  **(.83-15.9)** | **5.2**  **(1.7-15.5)** | **5.9**  **(1.7-20.7)** | **13.1**  **(7.9-21.8)** |
|  | 3 | **1.1**  **(.34-3.6)** | **1.3**  **(.32-5.6)** | **3.3**  **(1.2-9.6)** | **14.1**  **(7.0-28.1)** | **8.4**  **(5.0-14.1)** |
|  | 2 | **2.0**  **(1.2-3.4)** | **2.3**  **(1.0-5.2)** | **2.9**  **(1.1-7.6)** | **4.3**  **(1.6-11.3)** | **6.4**  **(2.7-15.0)** |
|  | 1 | **1.3**  **(.84-2.0)** | **4.4**  **(2.6-7.4)** | **3.0**  **(1.2-7.1)** | **2.7**  **(.82-9.2)** | **9.6**  **(4.0-23.0)** |
|  | 0 | **Ref** | **3.2**  **(1.9-5.3)** | **3.3**  **(1.4-8.0)** | **7.8**  **(2.5-23.9)** | **7.9**  **(2.2-28.8)** |
|  |  | 0 | 1 | 2 | 3 | 4+ |
| PEARLS | | | | | |  |

| Externalizing Problems | | | | | | |
| --- | --- | --- | --- | --- | --- | --- |
| QUIC | 4+ | **3.8**  **(.92-16.0)** | **0.0**  **(0.0)** | **0.0**  **(0.0)** | **3.3**  **(.44-24.8)** | **4.9**  **(1.8-13.9)** |
|  | 3 | **3.0**  **(1.5-6.1)** | **2.9**  **(.89-9.2)** | **2.8**  **(.68-11.7)** | **1.4**  **(.20-10.4)** | **7.0**  **(4.1-12.1)** |
|  | 2 | **2.0**  **(1.3-3.1)** | **4.5**  **(2.3-8.8)** | **1.7**  **(.41-6.9)** | **1.1**  **(.15-8.0)** | **6.6**  **(3.0-14.7)** |
|  | 1 | **1.5**  **(1.1-2.1)** | **1.8**  **(.94-3.5)** | **1.6**  **(.58-4.3)** | **4.5**  **(1.8-11.4)** | **2.9**  **(.90-9.4)** |
|  | 0 | **Ref** | **2.2**  **(1.4-3.4)** | **2.9**  **(1.5-5.8)** | **3.4**  **(1.2-9.4)** | **2.1**  **(.51-8.7)** |
|  |  | 0 | 1 | 2 | 3 | 4+ |
| PEARLS | | | | | |  |

| Externalizing Problems | | | | | | |
| --- | --- | --- | --- | --- | --- | --- |
| QUIC | 4+ | **0.0**  **(0.0)** | **11.6**  **(3.3-41.1)** | **13.5**  **(3.8-47.7)** | **0.0**  **(0.0)** | **6.4**  **(2.1-19.1)** |
|  | 3 | **2.0**  **(.60-7.0)** | **1.9**  **(.25-14.0)** | **2.4**  **(.31-18.0)** | **12.1**  **(4.0-36.8)** | **8.1**  **(3.2-20.6)** |
|  | 2 | **2.9**  **(1.4-5.9)** | **6.5**  **(2.8-15.0)** | **5.1**  **(1.5-17.5)** | **2.6**  **(.34-19.9)** | **6.6**  **(1.5-29.1)** |
|  | 1 | **1.3**  **(.65-2.6)** | **2.7**  **(1.0-7.4)** | **5.6**  **(1.6-19.2)** | **6.7**  **(1.5-29.9)** | **3.3**  **(.43-25.2)** |
|  | 0 | **Ref** | **1.4**  **(.41-4.7)** | **3.6**  **(.83-15.8)** | **9.2**  **(2.0-41.6)** | **0.0**  **(0.0)** |
|  |  | 0 | 1 | 2 | 3 | 4+ |
| PEARLS | | | | | |  |

| Sleep Disorders | | | | | | |
| --- | --- | --- | --- | --- | --- | --- |
| QUIC | 4+ | **2.5**  **(.88-7.1)** | **5.2**  **(2.1-12.7)** | **3.2**  **(1.1-9.3)** | **4.8**  **(1.6-14.2)** | **5.4**  **(3.2-9.2)** |
|  | 3 | **3.9**  **(2.4-6.4)** | **2.4**  **(1.0-5.7)** | **1.8**  **(.64-5.0)** | **5.7**  **(2.8-11.6)** | **4.1**  **(2.4-6.9)** |
|  | 2 | **2.2**  **(1.5-3.2)** | **2.0**  **(1.1-3.8)** | **2.0**  **(.84-4.6)** | **4.4**  **(2.0-9.4)** | **2.1**  **(.75-5.9)** |
|  | 1 | **1.3**  **(.93-1.8)** | **2.6**  **(1.6-4.1)** | **2.7**  **(1.3-5.6)** | **1.8**  **(.54-5.7)** | **1.3**  **(.31-5.5)** |
|  | 0 | **Ref** | **1.6**  **(.98-2.7)** | **1.7**  **(.68-4.3)** | **1.8**  **(.43-7.7)** | **0.0**  **(0.0)** |
|  |  | 0 | 1 | 2 | 3 | 4+ |
| PEARLS | | | | | |  |

| Sleep Disorders | | | | | | |
| --- | --- | --- | --- | --- | --- | --- |
| QUIC | 4+ | **2.1**  **(.82-5.3)** | **1.6**  **(.38-7.0)** | **3.4**  **(.98-11.7)** | **6.3**  **(2.6-15.0)** | **2.1**  **(1.0-4.4)** |
|  | 3 | **2.5**  **(1.7-3.7)** | **1.9**  **(.95-3.8)** | **2.6**  **(1.2-5.4)** | **1.7**  **(.67-4.2)** | **1.8**  **(1.2-3.0)** |
|  | 2 | **1.5**  **(1.2-2.0)** | **1.8**  **(1.1-3.0)** | **1.6**  **(.81-3.2)** | **2.7**  **(1.4-5.3)** | **2.7**  **(1.5-4.9)** |
|  | 1 | **1.4**  **(1.2-1.7)** | **2.1**  **(1.5-2.9)** | **1.5**  **(.92-2.5)** | **2.0**  **(1.0-3.8)** | **2.3**  **(1.2-4.4)** |
|  | 0 | **Ref** | **1.2**  **(.89-1.5)** | **2.1**  **(1.4-3.2)** | **2.0**  **(1.0-3.9)** | **2.2**  **(1.1-4.3)** |
|  |  | 0 | 1 | 2 | 3 | 4+ |
| PEARLS | | | | | |  |

| Abdominal Pain | | | | | | |
| --- | --- | --- | --- | --- | --- | --- |
| QUIC | 4+ | **2.5**  **(1.3-4.7)** | **.95**  **(.28-3.2)** | **.35**  **(.05-2.6)** | **1.4**  **(.53-3.7)** | **1.8**  **(1.1-3.1)** |
|  | 3 | **1.2**  **(.88-1.7)** | **1.6**  **(.94-2.6)** | **1.5**  **(.80-2.8)** | **1.2**  **(.58-2.4)** | **1.6**  **(1.1-2.2)** |
|  | 2 | **1.4**  **(1.2-1.7)** | **1.7**  **(1.2-2.4)** | **1.0**  **(.59-1.7)** | **1.3**  **(.71-2.4)** | **2.3**  **(1.4-3.5)** |
|  | 1 | **1.3**  **(1.2-1.5)** | **1.6**  **(1.3-2.1)** | **1.4**  **(1.0-2.0)** | **1.6**  **(.97-2.6)** | **2.2**  **(1.4-3.6)** |
|  | 0 | **Ref** | **1.1**  **(.93-1.4)** | **1.6**  **(1.2-2.2)** | **1.6**  **(.95-2.6)** | **1.6**  **(.93-2.7)** |
|  |  | 0 | 1 | 2 | 3 | 4+ |
| PEARLS | | | | | |  |

| Abdominal Pain | | | | | | |
| --- | --- | --- | --- | --- | --- | --- |
| QUIC | 4+ | 1.7  (.83-3.4) | **1.7**  **(.79-3.8)** | **2.1**  **(1.0-4.4)** | **3.1**  **(1.4-6.8)** | **2.0**  **(1.3-3.1)** |
|  | 3 | 1.4  (.95-2.1) | **2.1**  **(1.2-3.6)** | **1.3**  **(.71-2.5)** | **1.4**  **(.72-2.7)** | **1.7**  **(1.2-2.6)** |
|  | 2 | **1.8**  **(1.4-2.2)** | **2.1**  **(1.4-3.0)** | **2.4**  **(1.5-3.8)** | **1.1**  **(.55-2.3)** | **2.5**  **(1.4-4.4)** |
|  | 1 | **1.3**  **(1.1-1.6)** | **1.5**  **(1.1-2.0)** | **1.4**  **(.79-2.3)** | **2.3**  **(1.2-4.2)** | **1.8**  **(.90-3.7)** |
|  | 0 | **Ref** | **1.5**  **(1.1-2.1)** | **1.5**  **(.87-2.6)** | **1.4**  **(.57-3.4)** | **1.1**  **(.32-3.7)** |
|  |  | 0 | 1 | 2 | 3 | 4+ |
| PEARLS | | | | | |  |

| Asthma | | | | | | |
| --- | --- | --- | --- | --- | --- | --- |
| QUIC | 4+ | **1.7**  **(.88-3.4)** | **1.1**  **(.46-2.7)** | **1.1**  **(.48-2.8)** | **.77**  **(.23-2.6)** | **1.6**  **(1.0-2.5)** |
|  | 3 | **1.2**  **(.80-1.8)** | **1.2**  **(.67-2.3)** | **.86**  **(.41-1.8)** | **2.0**  **(1.1-3.6)** | **1.5**  **(1.0-2.4)** |
|  | 2 | **1.3**  **(1.1-1.7)** | **1.1**  **(.73-1.7)** | **1.5**  **(.88-2.5)** | **1.9**  **(1.0-3.5)** | **2.1**  **(1.1-3.9)** |
|  | 1 | **1.0**  **(.86-1.2)** | **.93**  **(.64-1.3)** | **2.0**  **(1.2-3.3)** | **1.4**  **(.66-2.8)** | **2.2**  **(1.1-4.3)** |
|  | 0 | **Ref** | **1.1**  **(.83-1.6)** | **2.1**  **(1.3-3.4)** | **1.6**  **(.71-3.8)** | **0.0**  **(0.0)** |
|  |  | 0 | 1 | 2 | 3 | 4+ |
| PEARLS | | | | | |  |

| Asthma | | | | | | |
| --- | --- | --- | --- | --- | --- | --- |
| QUIC | 4+ | **1.9**  **(.94-3.6)** | **1.2**  **(.42-3.7)** | **.72**  **(.17-3.1)** | **3.2**  **(1.4-7.1)** | **1.7**  **(.95-2.9)** |
|  | 3 | **1.4**  **(1.0-2.0)** | **1.0**  **(.56-1.8)** | **1.5**  **(.80-2.8)** | **.34**  **(.11-1.1)** | **1.8**  **(1.3-2.5)** |
|  | 2 | **1.3**  **(1.1-1.5)** | **1.1**  **(.76-1.7)** | **.99**  **(.57-1.7)** | **2.0**  **(1.1-3.3)** | **1.7**  **(1.0-2.8)** |
|  | 1 | **1.1**  **(.95-1.2)** | **1.4**  **(1.1-1.8)** | **1.4**  **(.98-2.0)** | **1.3**  **(.76-2.2)** | **1.7**  **(1.0-2.9)** |
|  | 0 | **Ref** | **1.1**  **(.90-1.3)** | **2.0**  **(1.5-2.7)** | **2.0**  **(1.3-3.2)** | **1.5**  **(.88-2.7)** |
|  |  | 0 | 1 | 2 | 3 | 4+ |
| PEARLS | | | | | |  |

| Headache | | | | | | |
| --- | --- | --- | --- | --- | --- | --- |
| QUIC | 4+ | **2.5**  **(1.3-5.1)** | **2.6**  **(1.2-5.8)** | **2.8**  **(1.3-5.9)** | **3.9**  **(1.7-8.9)** | **2.1**  **(1.3-3.3)** |
|  | 3 | **1.4**  **(.91-2.3)** | **1.8**  **(.98-3.5)** | **1.7**  **(.84-3.3)** | **1.9**  **(.95-3.8)** | **2.8**  **(1.8-4.1)** |
|  | 2 | **1.8**  **(1.4-2.3)** | **1.5**  **(.99-2.4)** | **1.0**  **(.51-2.1)** | **.86**  **(.34-2.2)** | **2.3**  **(1.2-4.5)** |
|  | 1 | **1.5**  **(1.2-1.8)** | **1.8**  **(1.3-2.5)** | **.99**  **(.49-2.0)** | **2.5**  **(1.3-4.9)** | **1.8**  **(.78-4.0)** |
|  | 0 | **Ref** | **1.6**  **(1.1-2.2)** | **2.8**  **(1.7-4.6)** | **1.3**  **(.47-3.8)** | **1.0**  **(.24-4.5)** |
|  |  | 0 | 1 | 2 | 3 | 4+ |
| PEARLS | | | | | |  |

| Headache | | | | | | |
| --- | --- | --- | --- | --- | --- | --- |
| QUIC | 4+ | **2.3**  **(1.1-4.9)** | **1.0**  **(.24-4.4)** | **2.0**  **(.57-7.0)** | **2.5**  **(.93-6.7)** | **2.2**  **(1.2-4.0)** |
|  | 3 | **1.7**  **(1.2-2.5)** | **1.5**  **(.83-2.9)** | **3.2**  **(1.7-6.0)** | **.63**  **(.20-2.0)** | **2.6**  **(1.8-3.7)** |
|  | 2 | **1.5**  **(1.2-1.9)** | **1.7**  **(1.1-2.6)** | **1.2**  **(.66-2.2)** | **1.7**  **(.85-3.3)** | **2.0**  **(1.2-3.5)** |
|  | 1 | **1.3**  **(1.1-1.5)** | **1.9**  **(1.4-2.4)** | **1.5**  **(1.0-2.3)** | **1.0**  **(.51-2.0)** | **1.7**  **(.90-3.1)** |
|  | 0 | **Ref** | **1.4**  **(1.1-1.7)** | **1.4**  **(.93-2.1)** | **1.2**  **(.57-2.3)** | **1.9**  **(1.0-3.6)** |
|  |  | 0 | 1 | 2 | 3 | 4+ |
| PEARLS | | | | | |  |

| Obesity | | | | | | |
| --- | --- | --- | --- | --- | --- | --- |
| QUIC | 4+ | **3.3**  **(1.9-5.9)** | **2.7**  **(1.1-6.2)** | **.94**  **(.35-2.5)** | **1.2**  **(.54-2.6)** | **1.5**  **(.97-2.4)** |
|  | 3 | **1.8**  **(1.4-2.3)** | **1.8**  **(1.2-2.7)** | **1.9**  **(1.2-3.2)** | **.73**  **(.40-1.3)** | **1.4**  **(1.1-1.9)** |
|  | 2 | **1.6**  **(1.4-1.9)** | **1.2**  **(.85-1.6)** | **1.4**  **(.95-2.0)** | **1.3**  **(.80-2.1)** | **1.5**  **(1.0-2.2)** |
|  | 1 | **1.3**  **(1.2-1.5)** | **1.2**  **(1.0-1.5)** | **1.1**  **(.81-1.4)** | **1.1**  **(.74-1.7)** | **2.0**  **(1.3-3.0)** |
|  | 0 | **Ref** | **.99**  **(.86-1.1)** | **.96**  **(.74-1.3)** | **1.1**  **(.69-1.6)** | **1.2**  **(.79-1.9)** |
|  |  | 0 | 1 | 2 | 3 | 4+ |
| PEARLS | | | | | |  |

| Obesity | | | | | | |
| --- | --- | --- | --- | --- | --- | --- |
| QUIC | 4+ | **2.6**  **(1.5-4.5)** | **1.7**  **(.91-3.2)** | **2.6**  **(1.4-4.8)** | **1.8**  **(.85-3.8)** | **2.0**  **(1.4-2.9)** |
|  | 3 | **1.7**  **(1.3-2.3)** | **1.5**  **(.97-2.4)** | **1.3**  **(.82-2.2)** | **1.5**  **(.87-2.4)** | **1.5**  **(1.1-2.1)** |
|  | 2 | **1.5**  **(1.3-1.8)** | **1.6**  **(1.2-2.2)** | **1.3**  **(.83-1.9)** | **1.2**  **(.69-2.0)** | **1.9**  **(1.2-3.2)** |
|  | 1 | **1.3**  **(1.1-1.5)** | **1.2**  **(.91-1.5)** | **1.2**  **(.79-1.8)** | **2.2**  **(1.3-3.8)** | **1.1**  **(.59-2.0)** |
|  | 0 | **Ref** | **1.0**  **(.82-1.3)** | **1.4**  **(.94-2.2)** | **1.8**  **(.92-3.6)** | **.91**  **(.35-2.4)** |
|  |  | 0 | 1 | 2 | 3 | 4+ |
| PEARLS | | | | | |  |
